# Case fatality rates for COVID-19 are higher than case fatality rates for motor vehicle accidents for individuals over 40 years of age

**DOI:** 10.1101/2021.04.09.21255193

**Authors:** Arjun Puranik, Michiel J.M. Niesen, Emily Lindemer, Patrick Lenehan, Tudor Cristea-Platon, Colin Pawlowski, Venky Soundararajan

## Abstract

The death toll of the COVID-19 pandemic has been unprecedented, due to both the high number of SARS-CoV-2 infections and the seriousness of the disease resulting from these infections. Here, we present mortality rates and case fatality rates for COVID-19 over the past year compared with other historic leading causes of death in the United States. Among the risk categories considered, COVID-19 is the third leading cause of death for individuals 40 years old and over, with an overall annual mortality rate of 325 deaths per 100K individuals, behind only cancer (385 deaths per 100K individuals) and heart disease (412 deaths per 100K individuals). In addition, for individuals 40 years old and over, the case fatality rate for COVID-19 is greater than the case fatality rate for motor vehicle accidents. In particular, for the age group 40-49, the relative case fatality rate of COVID-19 is 1.5 fold (95% CI: [1.3, 1.7]) that of a motor vehicle accident, demonstrating that SARS-CoV-2 infection may be significantly more dangerous than a car crash for this age group. For older adults, COVID-19 is even more dangerous, and the relative case fatality rate of COVID-19 is 29.4 fold (95% CI: [23.2, 35.7]) that of a motor vehicle accident for individuals over 80 years old. On the other hand, motor vehicle accidents have a 4.5 fold (95% CI: [3.9, 5.1]) greater relative case fatality rate compared to COVID-19 for the age group of 20-29 years. These results highlight the severity of the COVID-19 pandemic especially for adults above 40 years of age and underscore the need for large-scale preventative measures to mitigate risks for these populations. Given that FDA-authorized COVID-19 vaccines have now been validated by multiple studies for their outstanding real-world effectiveness and safety, vaccination of all individuals who are over 40 years of age is one of the most pressing public health priorities of our time.

## Introduction

As of April 1, 2021, over 550,000 deaths have been attributed to COVID-19 in the United States^1^, more than all of the deaths of American soldiers from the Vietnam war, Korean war, and World War II combined^2^. However, the disease burden of the pandemic has been largely unequal, and the majority of COVID-19 deaths have been concentrated among older adults (> 65 years old)^3^ and individuals with pre-existing medical conditions^4^. The CDC has identified these groups of individuals as “high risk” for severe COVID-19 disease^5^, and they have been prioritized in the nationwide COVID-19 vaccine rollout, along with healthcare and other essential workers^6^. Furthermore, increased mortality rates have also been observed among socioeconomically disadvantaged subgroups and racial minorities^3^.

Here, we assess the health risks and disease burden for COVID-19 and other leading causes of death in the United States over the past year, including: flu, cancer, heart disease, motor vehicle accidents, suicide, and homicide. We report the overall mortality rates and case fatality rates for 10-year age groups of the population, and we compute the relative risks for each of the non-COVID-19 causes of death versus COVID-19. For each age group, we present the absolute and relative risks of COVID-19. By placing the mortality risk of COVID-19 in the context of other prominent causes of death, within each age group, this analysis provides a useful point of reference for the danger of COVID-19.

## Methods

### Data Sources

Publicly available data was used for this analysis. Population data was obtained from the US Census Bureau^7^. National-level COVID-19 data was obtained from the Centers for Disease Control and Prevention (CDC) public use case surveillance data^8^. We restricted our analysis to the CDC-reported cases and deaths for the year spanning from March 15, 2020 to March 14, 2021. The CDC WONDER public access database was used to get overall mortality risks for a number of familiar causes of death, including heart disease and drug overdoses^9^. The mortality databases as well as cancer data were queried. In total, data from 4 of the datasets were used: *Cancer Incidence (2014-2017), Cancer Mortality 2014-2017), Detailed Mortality Underlying Cause of Death (2015-2018)*, and *Detailed Mortality Multiple Cause of Death (2015-2018)*. 5-year survival numbers for cancer were estimated from the Surveillance, Epidemiology and End Results (SEER) program data^10^ restricted to cases diagnosed from 2002-2016, along with life table data obtained from the US Mortality Database (USMDB)^11^. Overall mortality by motor vehicle accident was computed using the publicly available Fatality Analysis Reporting System (FARS) from the National Highway Traffic Safety Administration (NHTSA), which contains detailed information about all reported fatal accidents^12^. Case fatality rates (i.e. probability of dying conditioned on being in an accident) was derived from the publicly available NHTSA Crash Report Sampling System (CRSS)^13^. The Substance Abuse and Mental Health Services Administration (SAMHSA) National Survey on Drug Use and Health (NSDUH) was used to get survey-based age-specific estimates of annual drug misuse and misuse rates (including opioids and heroin) by age. Data was accessed through the survey’s *Public-use Data Analysis System*^*14*^. This was joined with the CDC WONDER drug fatality data, and the fatality numbers were divided by the estimated users to estimate the “conditional risk” of heroin use/opioid misuse. Note that the broadest category of the survey is used for the denominator, so frequent drug users likely have higher risk. Estimated deaths, illnesses, and hospitalizations by age group for influenza in the United States for the 2017-18 and 2018-2019 flu seasons were obtained from a recent CDC report^15,16^.

### Statistical analysis

For each age group and risk category, we computed the following measures:

- Prevalence = (Number of cases) / (Total population),
- Overall mortality = (Number of deaths) / (Total population),
- Case fatality rate = (Number of deaths) / (Number of cases).

For the estimates of the overall mortality and case fatality rates, individuals with missing or unknown death status were considered to be alive. The age group specific prevalence and overall mortality rate for COVID-19 were calculated by scaling up the number of cases and deaths captured by the CDC public use case surveillance dataset to the total number of cases and deaths reported in the CDC COVID data tracker^1^ during the study period (March 15, 2020 to March 14, 2021). Case fatality rates, along with standard errors, for motor vehicle accidents were computed using the survey package^17^ in R in order to account for the complex survey design. Case fatality rates for individuals in 4-door sedans using no restraint during an accident were computed using the same method. For cancer, 5-year mortality was used in place of the case fatality rate. In particular, the 5-year mortality rates for cancer along with standard errors were estimated from individuals who were diagnosed with cancer between 2002-2016 using the Ederer I method^18,19^. To compare the overall mortality and case fatality rates between other risks and COVID-19, we computed the following relative risks:

- Overall mortality relative risk = (Overall mortality for risk category) / (Overall mortality for COVID-19)
- Case fatality rate relative risk = (Case fatality rate for risk category) / (Case fatality rate for COVID-19)

In addition, 95% confidence intervals for the relative risks were computed using the delta method^20^. Rates were considered to be significantly different if the 95% confidence interval for the relative risk values did not contain one. This corresponds to a statistical significance test with p-value < 0.05.

We note that for influenza, estimates of case fatality and mortality data were only available for age groups of 0-4, 5-17, 18-49, 50-64, and 65+ years old.^16^ In addition, no confidence intervals were reported for the overall mortality and case fatality rate estimates for influenza because only uncertainty intervals were provided from the original data sources.

## Results

### COVID-19 is among the top-3 causes of death for individuals over 40 years old

Recently released statistics on the leading causes of death in the United States revealed that COVID-19 was the third most prevalent cause of death in 2020 (377,883 deaths, 11.3%), behind only Heart Disease and Cancer.^3^ We have stratified the mortality rates (deaths by cause per 100k individuals) of a diverse set of leading causes of death (defined in **Table S1**): Heart Disease, Cancer, COVID-19, Cerebrovascular Disease, Suicide, Motor Vehicle Accidents, Firearm Homicide, and Drug Overdoses, within each 10-year age interval. This analysis reveals that the high overall mortality of COVID-19 is mainly driven by high mortality rates for individuals over the age of 40 years (**Figure 1**). COVID-19 (red dots in **Figure 1**) is consistently among the top three causes of death in individuals aged 40 years or older. While COVID-19 ranks third among the causes of mortality in both the 40-49 age range as well as the 80+ age range, the overall mortality for the 40-49 age group is 21 deaths per 100K individuals, while it is 1,028 deaths per 100K individuals for the age group over 80 years old. These results show that most deaths from COVID-19 are among older age groups, which is consistent with previous studies^21,22^.

**Figure 1:**
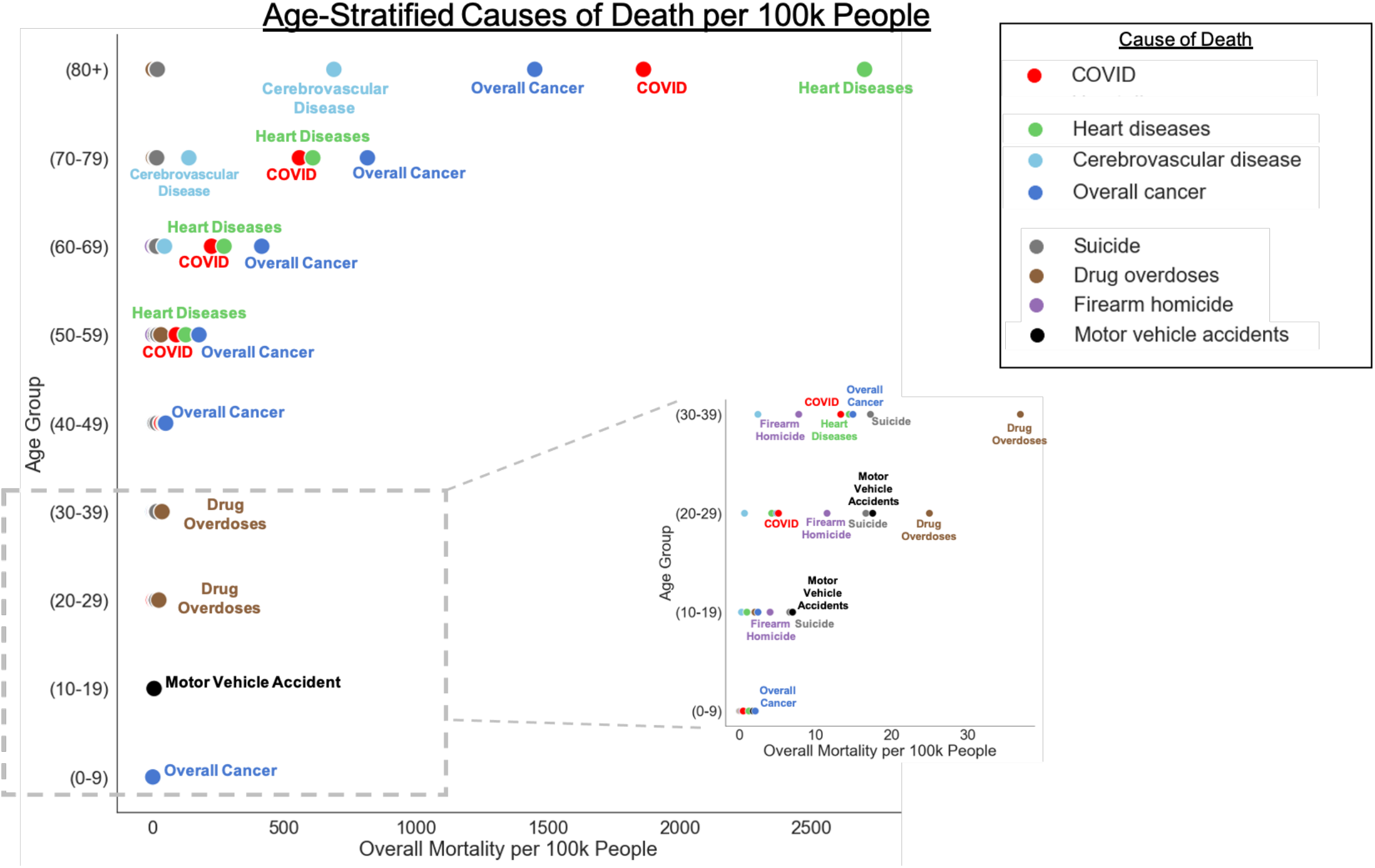
Mortality rates for leading causes of death in the United States stratified by age group. Comparison of causes of death per 100k people in a given age group, across all age groups. To better visualize mortality rates in the low-mortality age groups (ages 0 - 39), we have added a zoomed-in insert.

### Drug overdoses, suicide, and motor vehicle accidents are among the leading causes of death for individuals younger than 40 years old

For individuals under the age of 40, the leading causes of death are primarily environmental and mental health-related (**Figure 1**, inset). Drug overdoses are the leading cause of death for the age groups 30-39 and 20-29 (37 deaths and 25 deaths per 100K individuals, respectively). Suicide is the second leading cause of death for the age groups 10-19 and 30-39 (6.6 deaths and 17 deaths per 100K individuals, respectively), and third for the age group 20-29 (17 deaths per 100K individuals). Motor vehicle accidents are the leading cause of death for the age group 10-19 (7 deaths per 100K individuals) and the second-leading cause of death for the age group 20-29 (17 deaths per 100K individuals). For all age groups below 40 years old, the overall mortality rate of COVID-19 is low and it declines with each decade of age. There are 13 deaths per 100K individuals due to COVID-19 for the age group 30-39 years old, and for the age group 0-9 years old the overall mortality rate is less than one death per 100K individuals.

### COVID-19 has a higher case fatality rate than motor vehicle accidents for individuals over 40 years old

To further contextualize the risk of COVID-19 for individuals in varying age groups we analyzed the case fatality rate (CFR). This quantity is defined as the percentage of occurrences of a given event, such as a motor vehicle accident or contracting a disease, that result in death. Specifically, we compare the CFR of COVID-19 to that of motor vehicle accidents, specific cancers with high mortality,^23^ and the 2018-2019 flu (**Figure 2**). Strikingly, while for individuals in the younger age groups the CFR of motor vehicle accidents is significantly higher than that of COVID-19, the opposite is true for the older age groups. The largest difference, as quantified by the ratio of the motor vehicle accident CFR and the COVID-19 CFR, is observed between the 10-19 age group and the 80+ age group. Individuals in the 10-19 age group involved in a motor vehicle accident have a 13-fold (95% CI: [10, 17]) increased risk of death compared to individuals in the same age group who contract COVID-19 (**Table S2**). Conversely, individuals in the 80+ age group who contract COVID-19 have a 29-fold (95% CI: [24, 36]) increased risk of death compared to individuals in the same age group who are involved in a motor vehicle accident (**Table S2**). For individuals over 60 years old, the CFR of COVID-19 also surpasses that of prostate cancer (**Figure 2**), further emphasizing the dramatically increased risk of COVID-19 for older individuals. While the CFR for flu is also age dependent, it is significantly lower than the CFR for COVID-19 in comparable age-groups. Together, these data show that the risk of death upon contracting COVID-19 is greatly increased in older patients.

**Figure 2:**
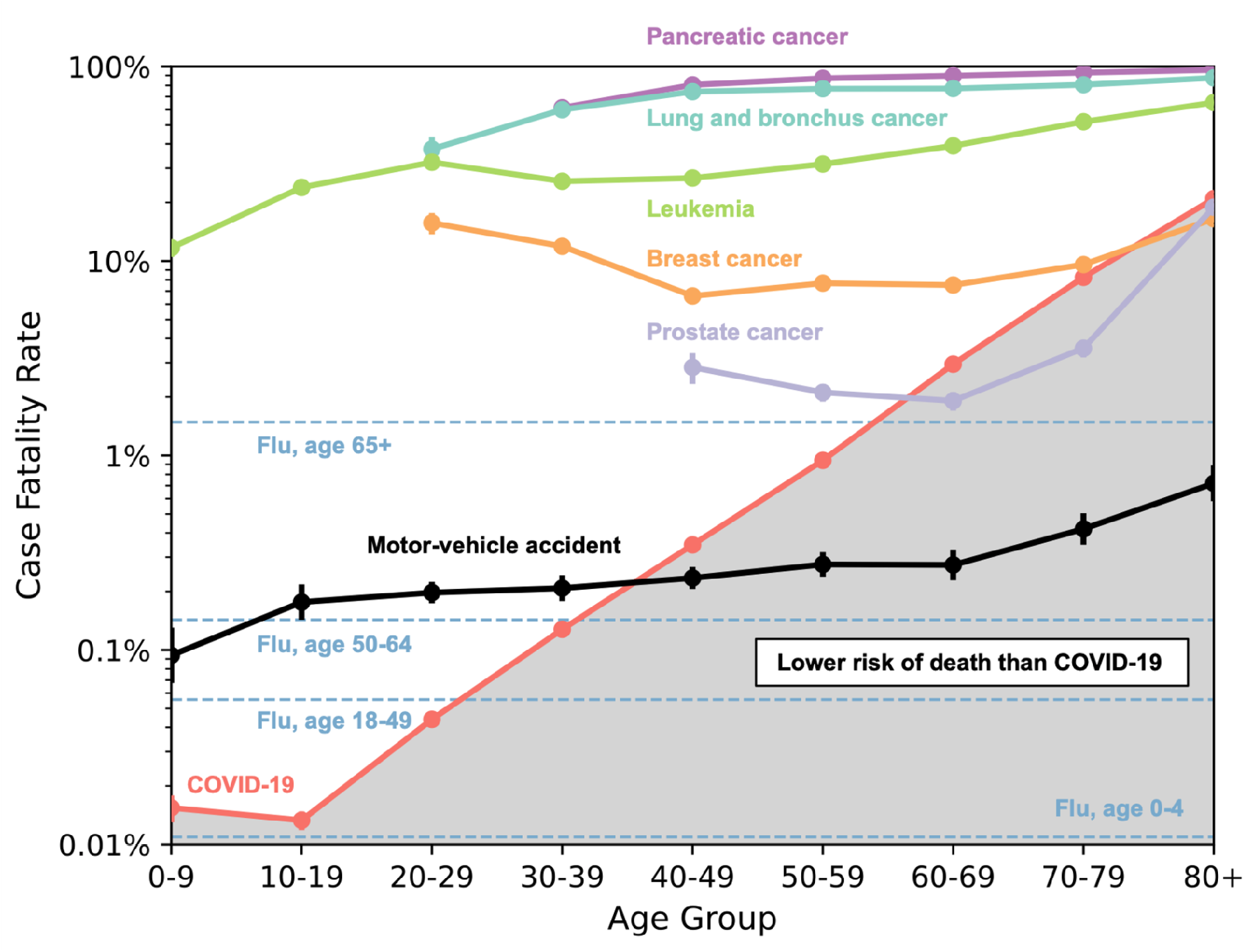
Case fatality rates for leading causes of death in the United States stratified by age group. Case fatality rate (CFR) per age group for COVID-19 (**red**), motor vehicle accidents (**black**), pancreatic cancer (**purple**), prostate cancer (**grey**), breast cancer (**orange**), lung cancer (**cyan**), leukemia (**green**), and the 2018-2019 flu (**blue**, horizontal dashed lines). Vertical bars show 95% confidence intervals, and the shaded region is a visual guide to indicate CFR values lower than COVID-19 CFR, per age group. Datapoints are not shown if a cause of death has a mortality of less than 0.1 deaths/100k individuals within an age group. Flu data is shown as horizontal lines due to flu data being stratified in different age groups^16^ than the other data shown. This data shows that for individuals in the age groups 40-49 years old and older, contracting COVID-19 results in a higher CFR than getting involved in a motor vehicle accident.

## Discussion

Here we have compared the mortality rate and case fatality rate of COVID-19 to other leading causes of death^3^, in the United States, stratified by 10-year age-groups (**Figure 1**). We find that COVID-19 mortality rates show a strong age dependence, with COVID-19 mortality increasing from less than 1 death per 100K individuals, for individuals under 20 years old, to 1,900 deaths per 100K individuals, for individuals over 80 years old. This increase in mortality can largely be attributed to high CFR in age groups over 40 years (**Figure 2**), where the risk of death for individuals with COVID-19 is greater than that of same aged individuals involved in a motor vehicle accident.

This study also corroborates previous studies on the leading causes of death in the United States. A recent study^24^ in December 2020 found that for the age group 85 years and older, the mortality rates for heart disease, COVID-19, and malignant neoplasms were 2,453 deaths, 1,070 deaths, and 1,044 deaths per 100K individuals respectively, which is similar to reported rates in this study for the age group 80 years and older (2,700 deaths, 1,900 deaths, and 1,500 deaths per 100K individuals, respectively). In addition, the case fatality rates for motor vehicle accidents reported here are consistent with data from the AAA Foundation’s American Driving Survey^25^, given that the motor vehicle accident CFR can be approximated as: (Number of Fatal Crashes) / (Total Number of Crashes). Although less pronounced than in the case of COVID-19, pancreatic cancer, lung and bronchus cancer, motor vehicle accidents, and the flu also show a near monotonic increase in CFR with age. On the other hand, breast cancer, prostate cancer, and leukemia have local maxima in CFR for younger age groups, consistent with previous studies that identified younger patients have poor prognosis for these types of cancer^26,27^.

There were a number of data challenges which limited the analysis in this study. During the first few months of 2020, counts of COVID-19 reported cases and deaths may be lower than the actual values due to underreporting. Case fatality rate data was sparse for the non-COVID-19 risk categories considered, so we focused the analysis on those categories with data available (e.g. motor vehicle accidents, cancer subtypes, influenza). Outcome measures for different risk categories were available for earlier time periods (e.g. cancer data was taken from 2002-2016), while COVID-19 outcome measures were taken from the past year. As a result, the outcome measures for the non-COVID-19 risk categories may be slightly different during the study time period from the past year. For example, the case fatality rates for cancer may be overestimated due to advances in cancer treatment^28^.

There are multiple areas for future work. Here, we consider COVID-19 overall mortality and case fatality rates as fixed, but it would be interesting to see how these outcome measures have changed over time. For example, we may expect that the mortality rates for COVID-19 have declined as therapeutics such as antiviral treatments^29^ and monoclonal antibodies^30,31^ have become available. As more follow-up data becomes available, we can report mortality rates for COVID-19 for longer time horizons. For example, in several years, we could conduct a follow-up study comparing the 5 year mortality rates from long-COVID with the 5 year mortality rates from cancer. In addition, one limitation of this study is that we restricted the analysis to mortality rates in the United States only. It would be interesting to replicate this analysis on data from developing countries which may have significantly different leading causes of death and mortality rates for COVID-19 and other risk categories.

## Data Availability

All data analyzed is publicly available freely and described in the Methods section

## Acknowledgments

The authors thank Murali Aravamudan and AJ Venkatakrishnan for their helpful feedback.

## Supplementary Materials

**Supplementary Table 1.** Risks considered and corresponding definitions of prevalence, risk-of-death, and mortality. Each risk measure is computed for each age group.

| Risk | Measure | Definition |
| --- | --- | --- |
| COVID-19 | Prevalence | [COVID cases reported to CDC] / [total US population] |
| COVID-19 | Risk-of-death | [COVID deaths] / [COVID cases] |
| COVID-19 | Overall mortality | [COVID deaths reported to CDC] / [US population] |
| Motor vehicle accidents | Prevalence | [estimated number of persons in motor vehicle accidents in the US based on CRSS, summed 2016-2018] / [total US population, summed 2016-2018] |
| Motor vehicle accidents | Risk-of-death | [total reported motor vehicle fatalities, summed 2016-2018] / [estimated number of persons in motor vehicle accidents in the US based on CRSS, summed 2016-2018] |
| Motor vehicle accidents | Overall mortality | [total reported motor vehicle fatalities, summed 2016-2018] / [total US population summed 2016-2018] |
| 4-door sedan accidents, with no seatbelt | Risk-of-death | [estimated number of persons who died in motor vehicle accidents while in a 4-door sedan wearing no seatbelt, from CRSS, summed 2016-2018] / [total estimated number of persons who were in motor vehicle accidents while in a 4-door sedan wearing no seatbelt, from CRSS, summed 2016-2018] |
| Flu in a given season | Prevalence | [CDC-estimated influenza medical visits in the US in the season] / [total US population] |
| Flu in a given season | Risk-of-death | [CDC-estimated influenza deaths in the US in the season] / [CDC-estimated influenza medical visits in the US in the season] |
| Flu in a given season | Overall mortality | [CDC-estimated influenza deaths in the US in the season] / [total US population] |
| Cancer, at a certain site | Prevalence | [CDC-reported total cancer incidence, summed 2015-2018] / [US population, summed 2015-2018] |
| Cancer, at a certain site | Risk-of-death | SEER data based calculation of relative mortality 5 years from diagnosis |
| Cancer, at a certain site | Overall mortality | [CDC-reported total cancer mortality, summed 2015-2018] / [US population, summed 2015-2018] |
| Opioid-involved overdose | Prevalence | [SAMHSA self-reported survey-based estimate of number of persons who misused opioids, summed 2015-2018] / [US population, summed 2015-2018] |
| Opioid-involved overdose | Risk-of-death | [CDC-reported number of opioid-involved drug overdoses] / [SAMHSA self-reported survey-based estimate of number of persons who misused opioids, summed 2015-2018] |
| Opioid-involved overdose | Overall mortality | [CDC-reported number of opioid-involved drug overdoses] / [US population, summed 2015-2018] |
| Heart disease, suicide, homicide, cerebrovascular disease, and other common causes of death | Overall mortality | [CDC-reported number of deaths from this cause, 2015-2018] / [US population, summed 2015-2018]<br>For these causes, we did not obtain additional data in order to estimate prevalence or risk-of-death, but we did include their more easily available overall mortality measure. |

**Supplementary Table 2:** Case fatality risk and overall mortality for leading causes of death stratified by age group, along with relative risks compared to COVID-19. For each age group, the top-5 leading causes of death are highlighted in red, and the rows showing the baseline rates for COVID-19 are in bold. The rows are sorted by age and overall mortality. All values are reported with two significant digits. The columns are: **(1) Age group:** 10-year age group considered, **(2) Risk category:** Cause of death considered, **(3) Prevalence per 100k:** Number of events per 100,000 individuals, **(4) Overall Mortality per 100k:** Number of deaths per 100,000 individuals, **(5) Case Fatality Rate:** Risk of death for individuals in the age group who experience the event, **(6) Overall Mortality Relative Risk:** (Overall Mortality for risk category) / (Overall Mortality for COVID-19) for the age group, along with 95% confidence interval, **(7) CFR Relative Risk:** (Case Fatality Risk for risk category) / (Case Fatality Risk for COVID-19) for the age group, along with the 95% confidence interval.

| Age group | Risk category | Prevalence per 100k | Overall Mortality per 100k | Case Fatality Risk | Overall Mortality Relative Risk (COVID-19 baseline) [95% CI] | CFR Relative Risk (COVID-19 baseline) [95% CI] |
| --- | --- | --- | --- | --- | --- | --- |
| 0-9 years | Overall cancer | 18 | 2.1 | 15% | 4.2 (3.6, 5.0) | 960 (820, 1100) |
| 0-9 years | Motor vehicle accidents | 2300 | 1.8 | 0.094% | 3.7 (3.1, 4.4) | 6.1 (4.2, 8.8) |
| 0-9 years | Heart diseases |  | 1.2 |  | 2.5 (2.1, 3.0) | N/A |
| 0-9 years | Brain and Other Nervous System cancer | 4 | 0.73 | 24% | 1.5 (1.3, 1.8) | 1500 (1300, 1800) |
| 0-9 years | Leukemias | 6 | 0.54 | 12% | 1.1 (0.93, 1.3) | 760 (630, 910) |
| **0-9 years** | **COVID-19** | **3300** | **0.49** | **0.015%** | **1** | **1** |
| 0-9 years | Cerebrovascular disease |  | 0.48 |  | 0.98 (0.82, 1.2) | N/A |
| 0-9 years | Firearm homicide |  | 0.32 |  | 0.66 (0.55, 0.79) | N/A |
| 0-9 years | Drug overdoses |  | 0.16 |  | 0.32 (0.26, 0.39) | N/A |
| 0-9 years | Lymphomas | 1.1 | 0.04 | 10.0% | 0.082 (0.062, 0.11) | 680 (490, 930) |
| 0-9 years | Suicide |  | 0.016 |  | 0.032 (0.021, 0.048) | N/A |
| 0-9 years | Lung and Bronchus cancer | 0.07 | 0.01 | 15% | N/A | 950 (320, 2900) |
| 0-9 years | Pancreas cancer | 0.02 | <0.01 | 37% | N/A | 2400 (780, 7500) |
| 0-9 years | Prostate cancer | 0.02 | <0.02 | 36% | N/A | 2300 (900, 5900) |
| 0-9 years | Female Breast cancer | 0 | 0 |  | N/A | N/A |
| 0-9 years | Motor vehicle accidents (4-door, no restraint) |  |  | 1.0% | N/A | 66 (26, 160) |
| 10-19 years | Motor vehicle accidents | 5200 | 7 | 0.18% | 7.2 (6.5, 8.1) | 13 (10.0, 17) |
| 10-19 years | Suicide |  | 6.6 |  | 6.8 (6.1, 7.7) | N/A |
| 10-19 years | Firearm homicide |  | 4 |  | 4.2 (3.7, 4.7) | N/A |
| 10-19 years | Overall cancer | 20 | 2.4 | 14% | 2.5 (2.3, 2.8) | 1000 (910, 1200) |
| 10-19 years | Drug overdoses |  | 2 |  | 2.1 (1.9, 2.4) | N/A |
| 10-19 years | Heart diseases |  | 0.97 |  | 1.0 (0.89, 1.1) | N/A |
| **10-19 years** | **COVID-19** | **7500** | **0.96** | **0.013%** | **1** | **1** |
| 10-19 years | Leukemias | 3.2 | 0.62 | 24% | 0.64 (0.56, 0.73) | 1800 (1600, 2100) |
| 10-19 years | Brain and Other Nervous System cancer | 2.6 | 0.59 | 16% | 0.62 (0.54, 0.7) | 1200 (1000, 1400) |
| 10-19 years | Cerebrovascular disease |  | 0.27 |  | 0.28 (0.24, 0.32) | N/A |
| 10-19 years | Lymphomas | 3.9 | 0.14 | 6.5% | 0.14 (0.12, 0.17) | 490 (400, 610) |
| 10-19 years | Lung and Bronchus cancer | 0.08 | 0.021 | 19% | 0.022 (0.015, 0.031) | 1400 (800, 2600) |
| 10-19 years | Pancreas cancer | 0.11 | <0.01 | 12% | N/A | 920 (310, 2800) |
| 10-19 years | Prostate cancer | <0.02 | <0.02 | 63% | N/A | 4800 (3000, 7600) |
| 10-19 years | Female Breast cancer | 0.15 | 0 | 6.3% | 0 [N/A] | 470 (120, 1900) |
| 10-19 years | Motor vehicle accidents (4-door, no restraint) |  |  | 2.8% | N/A | 210 (110, 380) |
| 20-29 years | Drug overdoses |  | 25 |  | 4.9 (4.7, 5.1) | N/A |
| 20-29 years | Motor vehicle accidents | 8600 | 17 | 0.2% | 3.4 (3.3, 3.6) | 4.5 (3.9, 5.1) |
| 20-29 years | Suicide |  | 17 |  | 3.2 (3.1, 3.4) | N/A |
| 20-29 years | Firearm homicide |  | 12 |  | 2.3 (2.1, 2.4) | N/A |
| **20-29 years** | **COVID-19** | **12000** | **5.1** | **0.044%** | **1** | **1** |
| 20-29 years | Overall cancer | 49 | 4.9 | 11% | 0.96 (0.91, 1.0) | 260 (240, 270) |
| 20-29 years | Heart diseases |  | 4.2 |  | 0.83 (0.79, 0.87) | N/A |
| 20-29 years | Leukemias | 2.9 | 0.86 | 32% | 0.17 (0.16, 0.18) | 730 (670, 800) |
| 20-29 years | Cerebrovascular disease |  | 0.66 |  | 0.13 (0.12, 0.14) | N/A |
| 20-29 years | Brain and Other Nervous System cancer | 2.5 | 0.61 | 19% | 0.12 (0.11, 0.13) | 430 (390, 480) |
| 20-29 years | Female Breast cancer | 6 | 0.42 | 16% | 0.082 (0.074, 0.092) | 360 (310, 410) |
| 20-29 years | Lymphomas | 7 | 0.39 | 9.6% | 0.077 (0.071, 0.084) | 220 (200, 250) |
| 20-29 years | Lung and Bronchus cancer | 0.42 | 0.11 | 38% | 0.021 (0.018, 0.024) | 860 (730, 1000) |
| 20-29 years | Pancreas cancer | 0.27 | 0.046 | 38% | 0.009 (0.0072, 0.011) | 850 (640, 1100) |
| 20-29 years | Prostate cancer | 0.02 | <0.02 | 37% | N/A | 850 (270, 2600) |
| 20-29 years | Motor vehicle accidents (4-door, no restraint) |  |  | 3.0% | N/A | 68 (53, 86) |
| 30-39 years | Drug overdoses |  | 37 |  | 2.8 (2.7, 2.9) | N/A |
| 30-39 years | Suicide |  | 17 |  | 1.3 (1.3, 1.3) | N/A |
| 30-39 years | Overall cancer | 120 | 15 | 13% | 1.1 (1.1, 1.2) | 100 (100, 110) |
| 30-39 years | Heart diseases |  | 14 |  | 1.1 (1.1, 1.1) | N/A |
| **30-39 years** | **COVID-19** | **11000** | **13** | **0.13%** | **1** | **1** |
| 30-39 years | Motor vehicle accidents | 6300 | 13 | 0.21% | 1.0 (0.97, 1.0) | 1.6 (1.4, 1.9) |
| 30-39 years | Firearm homicide |  | 7.8 |  | 0.59 (0.57, 0.61) | N/A |
| 30-39 years | Female Breast cancer | 46 | 4.7 | 12% | 0.35 (0.34, 0.37) | 93 (88, 99) |
| 30-39 years | Cerebrovascular disease |  | 2.4 |  | 0.18 (0.17, 0.19) | N/A |
| 30-39 years | Brain and Other Nervous System cancer | 3.8 | 1.4 | 23% | 0.11 (0.1, 0.11) | 180 (160, 190) |
| 30-39 years | Leukemias | 4.1 | 1.1 | 26% | 0.081 (0.076, 0.085) | 200 (190, 220) |
| 30-39 years | Lung and Bronchus cancer | 1.9 | 0.8 | 60% | 0.06 (0.057, 0.064) | 470 (440, 500) |
| 30-39 years | Lymphomas | 9 | 0.71 | 13% | 0.053 (0.05, 0.057) | 100 (92, 110) |
| 30-39 years | Pancreas cancer | 1.1 | 0.43 | 61% | 0.032 (0.03, 0.035) | 480 (440, 520) |
| 30-39 years | Prostate cancer | 0.37 | <0.02 | 11% | N/A | 84 (52, 130) |
| 30-39 years | Motor vehicle accidents (4-door, no restraint) |  |  | 3.6% | N/A | 28 (21, 38) |
| 40-49 years | Overall cancer | 320 | 51 | 18% | 1.4 (1.4, 1.4) | 51 (50, 52) |
| 40-49 years | Heart diseases |  | 46 |  | 1.3 (1.2, 1.3) | N/A |
| **40-49 years** | **COVID-19** | **11000** | **36** | **0.35%** | **1** | **1** |
| 40-49 years | Drug overdoses |  | 34 |  | 0.93 (0.91, 0.95) | N/A |
| 40-49 years | Suicide |  | 19 |  | 0.52 (0.51, 0.53) | N/A |
| 40-49 years | Female Breast cancer | 160 | 15 | 6.6% | 0.43 (0.42, 0.44) | 19 (18, 20) |
| 40-49 years | Motor vehicle accidents | 5500 | 12 | 0.23% | 0.34 (0.33, 0.35) | 0.68 (0.59, 0.77) |
| 40-49 years | Cerebrovascular disease |  | 7.6 |  | 0.21 (0.2, 0.22) | N/A |
| 40-49 years | Lung and Bronchus cancer | 13 | 6.9 | 74% | 0.19 (0.19, 0.2) | 210 (210, 220) |
| 40-49 years | Firearm homicide |  | 4.4 |  | 0.12 (0.12, 0.13) | N/A |
| 40-49 years | Brain and Other Nervous System cancer | 4.9 | 3 | 35% | 0.082 (0.079, 0.085) | 100 (97, 110) |
| 40-49 years | Pancreas cancer | 4.7 | 2.8 | 80% | 0.077 (0.074, 0.079) | 230 (230, 240) |
| 40-49 years | Leukemias | 7.5 | 1.9 | 27% | 0.052 (0.05, 0.054) | 77 (73, 80) |
| 40-49 years | Lymphomas | 15 | 1.6 | 18% | 0.044 (0.042, 0.046) | 52 (49, 55) |
| 40-49 years | Prostate cancer | 19 | 0.43 | 2.8% | 0.012 (0.011, 0.013) | 8.2 (6.8, 9.8) |
| 40-49 years | Motor vehicle accidents (4-door, no restraint) |  |  | 2.8% | N/A | 8.1 (5.3, 12) |
| 50-59 years | Overall cancer | 740 | 180 | 24% | 1.9 (1.9, 2.0) | 25 (25, 26) |
| 50-59 years | Heart diseases |  | 130 |  | 1.4 (1.4, 1.4) | N/A |
| **50-59 years** | **COVID-19** | **10000** | **91** | **0.94%** | **1** | **1** |
| 50-59 years | Lung and Bronchus cancer | 74 | 44 | 77% | 0.48 (0.47, 0.49) | 81 (80, 82) |
| 50-59 years | Female Breast cancer | 250 | 33 | 7.7% | 0.36 (0.36, 0.37) | 8.1 (7.9, 8.4) |
| 50-59 years | Drug overdoses |  | 33 |  | 0.36 (0.36, 0.37) | N/A |
| 50-59 years | Suicide |  | 21 |  | 0.23 (0.22, 0.23) | N/A |
| 50-59 years | Cerebrovascular disease |  | 20 |  | 0.21 (0.21, 0.22) | N/A |
| 50-59 years | Motor vehicle accidents | 4700 | 13 | 0.28% | 0.14 (0.14, 0.15) | 0.29 (0.25, 0.34) |
| 50-59 years | Pancreas cancer | 17 | 12 | 87% | 0.13 (0.13, 0.14) | 92 (91, 94) |
| 50-59 years | Brain and Other Nervous System cancer | 8.8 | 6.9 | 49% | 0.076 (0.074, 0.077) | 52 (51, 54) |
| 50-59 years | Prostate cancer | 170 | 5.7 | 2.1% | 0.063 (0.061, 0.065) | 2.2 (2.0, 2.5) |
| 50-59 years | Lymphomas | 29 | 4.4 | 23% | 0.048 (0.047, 0.049) | 24 (23, 25) |
| 50-59 years | Leukemias | 16 | 4.3 | 32% | 0.048 (0.047, 0.049) | 33 (32, 34) |
| 50-59 years | Firearm homicide |  | 2.5 |  | 0.027 (0.026, 0.028) | N/A |
| 50-59 years | Motor vehicle accidents (4-door, no restraint) |  |  | 4.6% | N/A | 4.8 (3.2, 7.4) |
| 60-69 years | Overall cancer | 1400 | 410 | 29% | 1.8 (1.8, 1.9) | 9.7 (9.6, 9.8) |
| 60-69 years | Heart diseases |  | 270 |  | 1.2 (1.2, 1.2) | N/A |
| **60-69 years** | **COVID-19** | **8000** | **220** | **2.9%** | **1** | **1** |
| 60-69 years | Lung and Bronchus cancer | 190 | 120 | 77% | 0.51 (0.51, 0.52) | 26 (26, 26) |
| 60-69 years | Female Breast cancer | 380 | 53 | 7.5% | 0.24 (0.23, 0.24) | 2.6 (2.5, 2.6) |
| 60-69 years | Cerebrovascular disease |  | 46 |  | 0.21 (0.2, 0.21) | N/A |
| 60-69 years | Pancreas cancer | 40 | 32 | 90% | 0.14 (0.14, 0.14) | 30 (30, 31) |
| 60-69 years | Prostate cancer | 490 | 29 | 1.9% | 0.13 (0.13, 0.13) | 0.65 (0.58, 0.72) |
| 60-69 years | Suicide |  | 17 |  | 0.075 (0.073, 0.076) | N/A |
| 60-69 years | Drug overdoses |  | 16 |  | 0.072 (0.071, 0.073) | N/A |
| 60-69 years | Brain and Other Nervous System cancer | 14 | 13 | 61% | 0.056 (0.055, 0.057) | 21 (20, 21) |
| 60-69 years | Motor vehicle accidents | 3600 | 12 | 0.27% | 0.053 (0.052, 0.054) | 0.093 (0.077, 0.11) |
| 60-69 years | Leukemias | 33 | 12 | 39% | 0.052 (0.051, 0.053) | 13 (13, 14) |
| 60-69 years | Lymphomas | 54 | 11 | 28% | 0.051 (0.05, 0.052) | 9.5 (9.2, 9.8) |
| 60-69 years | Firearm homicide |  | 1.4 |  | 0.006 (0.0057, 0.0063) | N/A |
| 60-69 years | Motor vehicle accidents (4-door, no restraint) |  |  | 1.6% | N/A | 0.53 (0.22, 1.3) |
| 70-79 years | Overall cancer | 2100 | 820 | 38% | 1.5 (1.5, 1.5) | 4.6 (4.6, 4.6) |
| 70-79 years | Heart diseases |  | 610 |  | 1.1 (1.1, 1.1) | N/A |
| **70-79 years** | **COVID-19** | **7100** | **560** | **8.2%** | **1** | **1** |
| 70-79 years | Lung and Bronchus cancer | 370 | 240 | 81% | 0.44 (0.44, 0.44) | 9.8 (9.7, 9.8) |
| 70-79 years | Cerebrovascular disease |  | 140 |  | 0.25 (0.25, 0.25) | N/A |
| 70-79 years | Prostate cancer | 580 | 91 | 3.6% | 0.16 (0.16, 0.17) | 0.43 (0.39, 0.48) |
| 70-79 years | Female Breast cancer | 460 | 82 | 9.6% | 0.15 (0.14, 0.15) | 1.2 (1.1, 1.2) |
| 70-79 years | Pancreas cancer | 70 | 61 | 93% | 0.11 (0.11, 0.11) | 11 (11, 11) |
| 70-79 years | Leukemias | 61 | 32 | 52% | 0.058 (0.057, 0.058) | 6.3 (6.2, 6.4) |
| 70-79 years | Lymphomas | 95 | 30 | 40% | 0.054 (0.053, 0.054) | 4.8 (4.7, 5.0) |
| 70-79 years | Brain and Other Nervous System cancer | 20 | 19 | 74% | 0.034 (0.033, 0.034) | 8.9 (8.8, 9.1) |
| 70-79 years | Suicide |  | 17 |  | 0.03 (0.029, 0.03) | N/A |
| 70-79 years | Motor vehicle accidents | 3000 | 13 | 0.42% | 0.023 (0.023, 0.024) | 0.051 (0.042, 0.061) |
| 70-79 years | Drug overdoses |  | 4.9 |  | 0.0088 (0.0085, 0.0091) | N/A |
| 70-79 years | Firearm homicide |  | 0.84 |  | 0.0015 (0.0014, 0.0016) | N/A |
| 70-79 years | Motor vehicle accidents (4-door, no restraint) |  |  | 6.5% | N/A | 0.79 (0.4, 1.5) |
| **80+ years** | **Heart diseases** |  | **2700** |  | **1.5 (1.4, 1.5)** | **N/A** |
| **80+ years** | **COVID-19** | **9300** | **1900** | **21%** | **1** | **1** |
| 80+ years | Overall cancer | 2200 | 1500 | 52% | 0.78 (0.78, 0.78) | 2.5 (2.5, 2.5) |
| 80+ years | Cerebrovascular disease |  | 690 |  | 0.37 (0.37, 0.37) | N/A |
| 80+ years | Lung and Bronchus cancer | 350 | 320 | 88% | 0.17 (0.17, 0.17) | 4.2 (4.2, 4.2) |
| 80+ years | Prostate cancer | 380 | 320 | 19% | 0.17 (0.17, 0.18) | 0.9 (0.85, 0.95) |
| 80+ years | Female Breast cancer | 360 | 150 | 17% | 0.079 (0.078, 0.08) | 0.79 (0.75, 0.83) |
| 80+ years | Pancreas cancer | 94 | 99 | 96% | 0.053 (0.053, 0.054) | 4.6 (4.6, 4.6) |
| 80+ years | Leukemias | 85 | 72 | 65% | 0.038 (0.038, 0.039) | 3.1 (3.1, 3.2) |
| 80+ years | Lymphomas | 110 | 67 | 58% | 0.036 (0.036, 0.037) | 2.8 (2.7, 2.8) |
| 80+ years | Brain and Other Nervous System cancer | 20 | 21 | 82% | 0.011 (0.011, 0.012) | 3.9 (3.8, 4.0) |
| 80+ years | Suicide |  | 19 |  | 0.01 (0.01, 0.01) | N/A |
| 80+ years | Motor vehicle accidents | 2100 | 17 | 0.72% | 0.0094 (0.0091, 0.0096) | 0.034 (0.028, 0.042) |
| 80+ years | Drug overdoses |  | 3.7 |  | 0.002 (0.0019, 0.0021) | N/A |
| 80+ years | Firearm homicide |  | 0.65 |  | 0.00035 (0.00031, 0.00039) | N/A |
| 80+ years | Motor vehicle accidents (4-door, no restraint) |  |  | 7.7% | N/A | 0.37 (0.16, 0.82) |

**Supplementary Table 3:** Estimated case fatality risk and overall mortality for flu stratified by age groups, along with estimated relative risks compared to COVID-19. This is a continuation of Supplementary Table 2 for the case of flu for the age groups 0-4, 5-17, 18-49, 50-64, and 65+ years old as reported by the CDC. In the first five rows, we provide data for the 2017-18 flu season, and in the last five rows we provide data for the 2018-19 flu season.^15,16^

| Age group | Risk category | Prevalence per 100k | Overall Mortality per 100k | Case Fatality Risk |
| --- | --- | --- | --- | --- |
| 0-4 years | Flu 2017-18 | 12000 | 0.58 | 0.0047% |
| 5-17 years | Flu 2017-18 | 7300 | 0.99 | 0.014% |
| 18-49 years | Flu 2017-18 | 3900 | 2 | 0.053% |
| 50-64 years | Flu 2017-18 | 9000 | 11 | 0.12% |
| 65+ years | Flu 2017-18 | 6400 | 97 | 1.5% |
| 0-4 years | Flu 2018-19 | 12000 | 1.4 | 0.011% |
| 5-17 years | Flu 2018-19 | 7500 | 0.39 | 0.0053% |
| 18-49 years | Flu 2018-19 | 3200 | 1.8 | 0.056% |
| 50-64 years | Flu 2018-19 | 6300 | 9 | 0.14% |
| 65+ years | Flu 2018-19 | 3200 | 47 | 1.5% |

## References

1. CDC. COVID Data Tracker. https://covid.cdc.gov/covid-data-tracker (2020).

2. McCarthy, N. U.S. Deaths From Covid-19 Match Toll Of Three Major Wars. <https://www.statista.com/chart/24252/us-covid-19-deaths-compared-to-deaths-in-major-wars/>.

3. Ahmad, F. B. Provisional Mortality Data — United States, 2020. MMWR Morb. Mortal. Wkly. Rep. 70, (2021).

4. Weekly Updates by Select Demographic and Geographic Characteristics. <https://www.cdc.gov/nchs/nvss/vsrr/covid_weekly/index.htm?fbclid=IwAR3-wrg3tTKK5-9tOHPGAHWFVO3DfslkJ0KsDEPQpWmPbKtp6EsoVV2Qs1Q#Comorbidities (2021).

5. CDC. Healthcare Workers. <https://www.cdc.gov/coronavirus/2019-ncov/hcp/clinical-care/underlying-evidence-table.html?CDC_AA_refVal=https%3A%2F%2Fwww.cdc.gov%2Fcoronavirus%2F2019-ncov%2Fneed-extra-precautions%2Fevidence-table.html (2021).

6. CDC. CDC’s COVID-19 Vaccine Rollout Recommendations. https://www.cdc.gov/coronavirus/2019-ncov/vaccines/recommendations.html (2021).

7. US Census Bureau. 2019 Population Estimates by Age, Sex, Race and Hispanic Origin.

8. Calgary, O. COVID-19 Case Surveillance Public Use Data. https://data.cdc.gov/Case-Surveillance/COVID-19-Case-Surveillance-Public-Use-Data/vbim-akqf/data.

9. CDC WONDER. https://wonder.cdc.gov/.

10. SEER Cancer Statistics Review, 1975-2016. https://seer.cancer.gov/csr/1975_2016/index.html.

11. United States Mortality Database. https://usa.mortality.org/.

12. Fatality Analysis Reporting System (FARS). <https://www.nhtsa.gov/research-data/fatality-analysis-reporting-system-fars> (2016).

13. Crash Report Sampling System. <https://www.nhtsa.gov/crash-data-systems/crash-report-sampling-system> (2017).

14. SAMHDA. https://pdas.samhsa.gov/#/.

15. Estimated Influenza Illnesses, Medical visits, Hospitalizations, and Deaths in the United States — 2017–2018 influenza season. <https://w>ww.cdc.gov/flu/about/burden/2017-2018.htm (2021).

16. Estimated Influenza Illnesses, Medical visits, Hospitalizations, and Deaths in the United States — 2018–2019 influenza season. <https://www.cdc.gov/flu/about/burden/2018-2019.html> (2021).

17. survey: Analysis of Complex Survey Samples. https://CRAN.R-project.org/package=survey.

18. Cho H, Howlader N, Mariotto AB, Cronin KA. Estimating relative survival for cancer patients from the SEER Program using expected rates based on Ederer I versus Ederer II method. Surveillance Research Program, National Cancer Institute; 2011. Technical Report #2011- Available from: https://surveillance.cancer.gov/reports/.

19. Ederer, F., Axtell, L. M. & Cutler, S. J. The relative survival rate: a statistical methodology. Natl. Cancer Inst. Monogr. 6, 101–121 (1961).

20. Fernandez, M. A. L., MA, MPH & MSc. Delta Method in Epidemiology: An Applied and Reproducible Tutorial. https://migariane.github.io/DeltaMethodEpiTutorial.nb.html.

21. Jordan, R. E., Adab, P. & Cheng, K. K. Covid-19: risk factors for severe disease and death. BMJ 368, (2020).

22. Selvan, M. E. Risk factors for death from COVID-19. Nat. Rev. Immunol. 20, 407–407 (2020).

23. CDCBreastCancer. An Update on Cancer Deaths in the United States. <https://www.cdc.gov/cancer/dcpc/research/update-on-cancer-deaths/index.htm >(2021).

24. Woolf, S. H., Chapman, D. A. & Lee, J. H. COVID-19 as the Leading Cause of Death in the United States. JAMA 325, 123–124 (2021).

25. Tefft, B. Rates of Motor Vehicle Crashes, Injuries and Deaths in Relation to Driver Age, United States, 2014-2015. AAA Foundation for Traffic Safety. (2017).

26. Collins, L. C. et al.. Pathologic features and molecular phenotype by patient age in a large cohort of young women with breast cancer. Breast Cancer Res. Treat. 131, 1061–1066 (2012).

27. Salinas, C. A., Tsodikov, A., Ishak-Howard, M. & Cooney, K. A. Prostate cancer in young men: an important clinical entity. Nat. Rev. Urol. 11, 317–323 (2014).

28. Siegel, R. L., Miller, K. D. & Jemal, A. Cancer statistics, 2020. CA Cancer J. Clin. 70, 7–30 (2020).

29. Beigel, J. H. et al.. Remdesivir for the Treatment of Covid-19 - Final Report. N. Engl. J. Med. 383, 1813–1826 (2020).

30. Mahase, E. Covid-19: FDA authorises neutralising antibody bamlanivimab for non-admitted patients. BMJ 371, (2020).

31. Cohen, M. S. Monoclonal Antibodies to Disrupt Progression of Early Covid-19 Infection. The New England journal of medicine vol. 384 289–291 (2021).

